# Suitability of Two Rapid Lateral Flow Immunochromatographic Assays for Predicting SARS-CoV-2 Neutralizing Activity of Sera

**DOI:** 10.1101/2020.09.23.20198713

**Authors:** Arantxa Valdivia, Ignacio Torres, Víctor Latorre, Clara Francés-Gómez, Josep Ferrer, Lorena Forqué, Rosa Costa, Carlos Solano de la Asunción, Dixie Huntley, Roberto Gozalbo-Rovira, Javier Buesa, Estela Giménez, Jesús Rodríguez-Díaz, Ron Geller, David Navarro

## Abstract

**Purpose:** Assessment of commercial SARS-CoV-2 immunoassays for their capacity to provide reliable information on sera neutralizing activity is an emerging need. We evaluated the performance of two commercially-available lateral flow immunochromatographic assays (LFIC) (Wondfo SARS-CoV-2 Antibody test and the INNOVITA 2019-nCoV Ab test) in comparison with a SARS-CoV-2 neutralization pseudotyped assay for COVID-19 diagnosis in hospitalized patients, and investigate whether the intensity of the test band in LFIC associates with neutralizing antibody (NtAb) titers.

**Patients and Methods:** Ninety sera were included from 51 patients with moderate to severe COVID-19. A green fluorescent protein (GFP) reporter-based pseudotyped neutralization assay (vesicular stomatitis virus coated with SARS-CoV-2 spike protein) was used. Test line intensity was scored using a 4-level scale (0 to 3+).

**Results:** Overall sensitivity of LFIC assays was 91.1% for the Wondfo SARS-CoV-2 Antibody test, 72.2% for the INNOVITA 2019-nCoV IgG, 85.6% for the INNOVITA 2019-nCoV IgM and 92.2% for the NtAb assay. Sensitivity increased for all assays in sera collected beyond day 14 after symptoms onset (93.9%, 79.6%,93.9% and 93.9%, respectively). Reactivities equal to or more intense than the positive control line (≥2+) in the Wondfo assay had a negative predictive value of 100% and a positive predictive value of 96.4% for high NtAb_50_ titers (≥1/160).

**Conclusions:** Our findings support the use of LFIC assays evaluated herein, particularly the Wondfo test, for COVID-19 diagnosis. We also find evidence that these rapid immunoassays can be used to predict high SARS-CoV-2-S NtAb_50_ titers.

## Introduction

Serological testing is increasingly recognized as a useful tool for control of the COVID-19 pandemic; beyond complementing RT-PCR assays for disease diagnosis in symptomatic patients, detection of specific antibodies allows for estimating SARS-CoV-2 infection incidence and virus spread in a given population, infering protection against reinfection, evaluating vaccine efficacy and selecting appropriate plasma specimens from convalescent COVID-19 patients for passive transfer therapies [1-3]. Numerous SARS-CoV-2 serological tests have been commercialized [4], among which lateral flow immunochromatographic assays (LFIC) are particularly appealing because of their rapid turnaround times, simplicity of use and suitability for point of care testing.

SARS-CoV-2 neutralizing antibodies (NtAb) are presumed to play a major protective role against SARS-CoV-2 infection [5-7]. Unfortunately, virus neutralization assays, whether using wild-type SARS-CoV-2, engineered SARS-CoV-2 pseudotypes or chimeric viruses [8] are unsuited for routine testing, thus creating a need to assess commercial SARS-CoV-2 immunoassays for their capacity to provide reliable information on sera neutralizing activity. Several studies have evaluated the performance of LFIC in comparison with NtAb assays in subjects with past or ongoing SARS-CoV-2 infection [9-12]. Nevertheless, to the best of our knowledge, only one has attempted to quantitatively correlate results yielded by the two assay types by analyzing the strength of test line reactivity in LFIC devices and NtAb_50_ titers [11].

Here, we sought to evaluate the performance of two commercially-available LFIC, widely used in our country, compared with a SARS-CoV-2 neutralization pseudotype assay for COVID-19 diagnosis in hospitalized patients, and determine whether the intensity of the test band in LFIC was associated with levels of NtAb recognizing the SARS-CoV-2 Spike (S) protein.

## Material and Methods

### Serum specimens and patients

The current study included 90 sera from 51 patients with moderate to severe laboratory-confirmed (RT-PCR) COVID-19 RT-PCR [13] admitted to Hospital Clínico Universitario of Valencia between March 5 and April 30, 2020. Sera were grouped according to the timing of collection after symptoms onset: 41 were obtained within < 15 days (median, 11 days; range, 5-14 days), and 49 later on (≥ 15 days, at a median of 23 days; range, 15-41 days). Sera had been cryopreserved at -20 °C and were thawed for the analyses described below. This study was approved by the Research Ethics Committee of Hospital Clínico Universitario INCLIVA (March, 2020).

### SARS-CoV-2 Neutralizing antibody assay

A green fluorescent protein (GFP) reporter-based pseudotyped neutralization assay with a non-replicative vesicular stomatitis virus (VSV) backbone coated with SARS-CoV-2 spike (S) protein was used for neutralization assays on Vero cells, using heat-inactivated sera and a viral input of 1,250 focus-forming units, as previously described [13]. Sera which did not reduce viral replication by 50% at 1/20 dilution were considered non-neutralizing and were arbitrarily assigned a value of 1/10. The antibody dilution resulting in 50% virus neutralization (NtAb_50_) was calculated using the drc package (version 3.0-1) in R via a two-parameter log-logistic regression model (LL.2 model).

### Commercial SARS-CoV-2 IgG LFIC immunoassays

Two LFIC were evaluated: SARS-COV-2 Antibody test from Guangzhou Wondfo Biotech Co., Ltd. (China), which detects SARS-CoV-2 antibodies (IgG and IgM) in a single test band, and INNOVITA 2019-nCoV Ab Test (Beijing Innovita Biological technology, China) which detects IgG and IgM separately. The antigenic specificity of antibodies detected by these assays was not disclosed (to our knowledge). Both assays were performed according to the protocol provided by the respective manufacturer. Test line intensity scoring was done using a 4-level scale (Figure 1), in which 0 corresponded to a negative result (absence of a test line), 1+ represented a weak positive result (intensity of test band lower than control band), 2+ a positive result (intensity of test band equal to control line), and 3+ a strong positive result (intensity of test band greater than control line). Four readers independently scored each test and the average of the scores was recorded as the final LFIC result.

**Figure 1.**
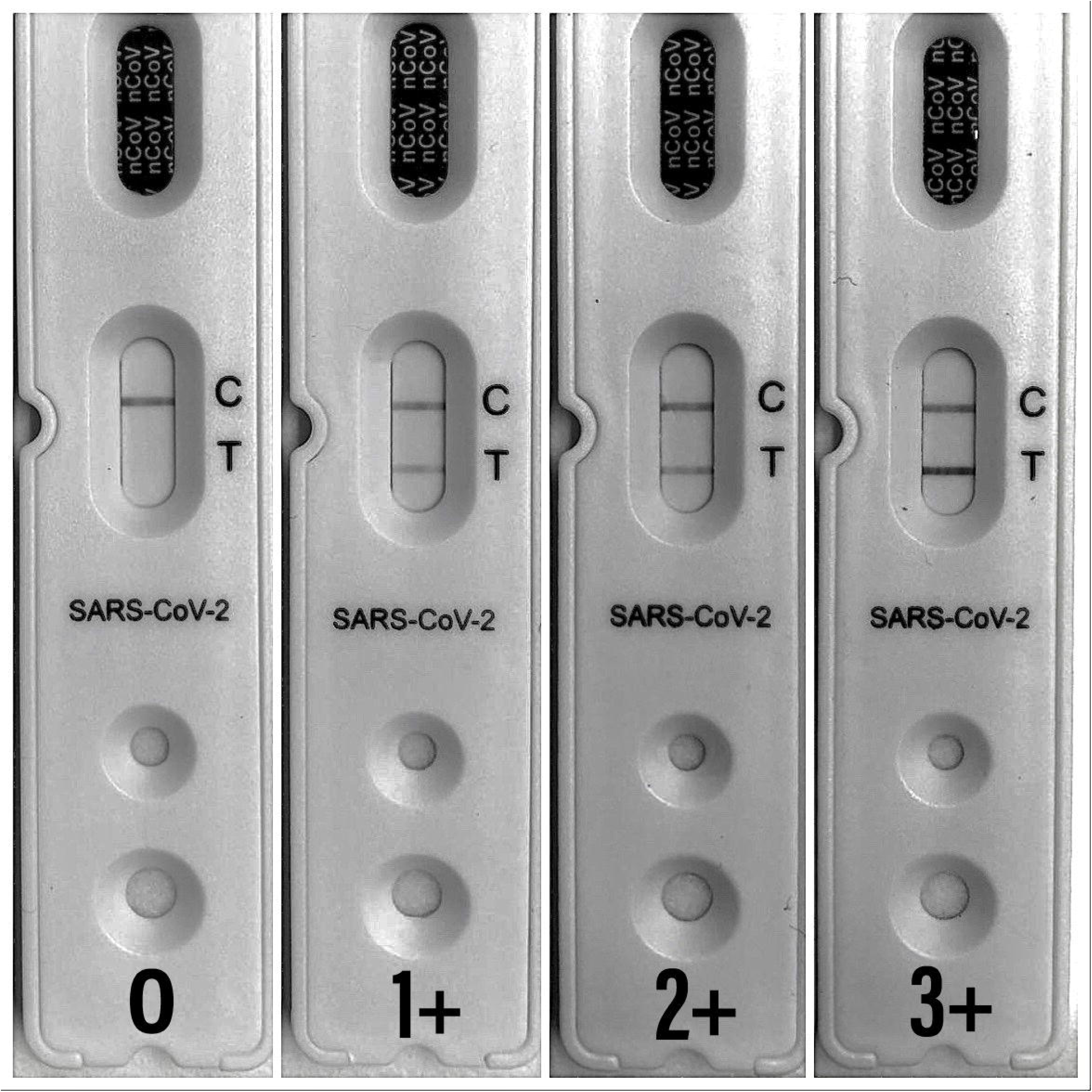
Test line intensity was scored using a 4-level scale. From left to right; 0: negative result, 1+: weak positive result (intensity of test band lower than the control band); 2+: positive result (intensity of test band equal to the control line); 3+: strong positive result (intensity of test band greater than the control line).

### Definition

Here, NtAb titers ≥1/160 were deemed as high, in line with the minimum NtAb_50_ titer of plasma from COVID-19 convalescent individuals recommended by FDA for therapeutic use [14].

### Statistical methods

Test performances were evaluated by sensitivity with the associated 95% confidence interval (CI). Cohen’s Kappa (κ) statistic was used to evaluate the qualitative agreement between immunoassays. Differences between medians were compared using the Mann-Whitney U-test. A *P*-value <0.05 was considered statistically significant. The analyses were performed using SPSS version 20.0 (SPSS, Chicago, IL, USA).

## Results

### LFIC immunoassays performance

Categorical results obtained by the LFIC immunoassays and the NtAb assay are shown in Table 2. Out of a total of 90 sera, 83 tested positive by the NtAb assay, 82 by Wondfo SARS-CoV-2 Antibody test, 65 by INNOVITA 2019-nCoV IgG, and 74 by INNOVITA 2019-nCoV IgM. Taking RT-PCR positive results as the reference, the data showed that the Wondfo SARS-COV-2 Antibody test had higher overall sensitivity than the INNOVITA 2019-nCoV IgG and IgM, considered either separately or in combination, and approached that of NtAb, the most sensitive assay (92.2%) (Table 2). As expected, the sensitivity of LFIC assays increased when testing sera collected at relatively late times (≥15 days) after symptoms onset compared with sera collected earlier (<15 days). Notably, the Wondfo SARS-CoV-2 Antibody test, INNOVITA 2019-nCoV IgM and NtAb assay showed similar sensitivity for late sera. Considering the entire data set, the degree of agreement between qualitative results yielded by the commercial immunoassays and the NtAb assay was κ=0.64 (*P*=<0.001) for Wondfo SARS-CoV-2 Antibody test, κ=0.29 (*P*=<0.001) for INNOVITA 2019-nCoV IgG, κ=0.46 (*P*=<0.001) for INNOVITA 2019-nCov IgM, and κ=0.56 (*P*=<0.001) for the combination of INNOVITA 2019-nCoV IgG and IgM testing.

### Prediction of high NtAb titers according to LFIC test line intensity

Median NtAb_50_ titers increased significantly in parallel with test line intensity (from 0 to 3+) in the Wondfo LFIC device (Figure 2). In contrast, median NtAb_50_ titers were not associated with strength of reactivity of the positive test line (1+ to 3+) in the INNOVITA LFIC assay, either for IgG or IgM, although they were significantly different in sera returning negative results (0) from those yielding positive results (1+ to 3+).

**Figure 2.** Median neutralizing antibody titers (NtAb_50_) in sera from hospitalized COVID-19 patients according to their strength of reactivity in lateral flow immunochromatographic assays (grading scale of test lines from 0 to 3+). *P* values for selected comparisons are shown.

High NtAb_50_ titers (≥1/160) were observed in 74 out of 90 sera. The Wondfo SARS-CoV-2 Antibody test best predicted NtAb_50_ titers ≥1/160, with reactivities ≥2+ having a negative predictive value (NPV) of 100% and a positive predictive value (PPV) of 96.4 (Table 3). In contrast, both the IgG and IgM INNOVITA LFIC assay exhibited good PPVs but suboptimal NPVs at a threshold of ≥2+. Setting the cut-off at a weaker band intensity (1+) returned slightly worse results for all LFIC

## Discussion

In this study the Wondfo SARS-CoV-2 Antibody LFIC assay was found to exhibit excellent overall sensitivity (91.1%) in a cohort COVID-19 patients with moderate to severe forms of the disease. As expected, sensitivity was higher for sera collected beyond day 14 after onset of symptoms than for those drawn earlier on. An even higher sensitivity for the Wondfo assay than that observed in the current study was reported by Martínez and colleagues [15] in a mixed cohort including SARS-CoV-2 infected asymptomatic individuals and patients presenting with mild to severe COVID-19. In turn, Guedes-López et al. [16] reported a sensitivity of 83% when testing sera collected between days 15-28 after symptoms onset in a cohort comprising symptomatic healthcare workers and patients admitted to the Emergency Department.

Overall, the INNOVITA 2019-nCoV assay performed worse than the Wondfo SARS-CoV-2 Antibody LFIC assay in terms of sensitivity (72.2% for IgG and 82.2% for IgM), although combining the results of both test lines yielded an acceptable figure (85.6%). Nevertheless, when only sera obtained late after symptoms onset were analyzed, sensitivity was similar to the Wondfo assay (93.9%). To our knowledge, only one study has evaluated the performance of the INNOVITA LFIC assay [17]. Yong et al. [17] reported a sensitivity of 50% and 52% for IgM, and 87.5% and 91.3% in sera collected within 8-15 days and ≥15 days after the onset of symptoms, respectively, in a cohort of hospitalized COVID-19 patients. Differences in the precise timing of sera collection and the severity of COVID-19 may account for these rather minor discrepancies across the above-mentioned studies; as for the latter, Martínez et al. [15] found a lower sensitivity for asymptomatic individuals (84.6%) than for the entire study group (89.9%).

A handful of studies have compared the performance of LFIC assays versus SARS-CoV-2 neutralization assays [9-12], but none of the LFIC evaluated herein were included. These studies differed in many aspects, namely, the timing of sera collection, clinical presentation of COVID-19, neutralization antibody assay employed, NtAb_50_ titer cutoff value for positive results, and the reference method for sensitivity calculations (either the NtAb assay itself or RT-PCR results). Not surprisingly, overall sensitivities reported for these LFIC assays vary widely, ranging from 46% [11] to 100%. [10]. Here we used a SARS-CoV-2-S-pseudotype as the viral input; nevertheless, NtAb levels measured by this assay have been shown to correlate strongly with those from assays using live SARS-CoV-2 [8]. Although the overall sensitivity of both LFIC assays was lower than the neutralization assay, it was comparable when sera collected ≥15 days were analyzed separately.

A novel contribution of the current study is that a strong association was found between the strength of test line reactivity and NtAb_50_ titers, notably when employing the Wondfo SARS-CoV-2 Antibody assay. Using a simple grading scale, we could discriminate reasonably well between sera containing high and low NtAb_50_ titers (≥1/160 or <1/160, respectively). In sera giving reactivities ≥2+ (comparable to or more intense than the control line) we identified high-NtAb level sera with excellent PPV and NPV. Weidner and colleagues [9] were the first to prove the suitability of this approach. By using a 4-level intensity scale on the Wantai SARS-CoV-2 Ab rapid test, they were able to predict NtAb_50_ titers (live SARS-CoV-2) >1/200 with NPV and PPV of 92%. The Wantai assay detects antibodies binding the S receptor binding domain which includes several highly immunogenic epitopes eliciting potent NtAb responses within several epitopes [18,19], making the above-reported association somewhat unsurprising. The antigenic target/s of the LFIC assays evaluated herein are unknown to us, thus precluding speculation on that matter.

The observer-dependent scoring of test line reactivity may be construed as a limitation of this study, although in fact all four readers evaluating LFIC results concurred in the categorization of all sera. Moreover, readings were consistent across different rounds of testing (not shown).

In summary, our data support the use of all LFIC assays evaluated herein, particularly the Wondfo test, for COVID-19 diagnosis, especially when testing sera collected late after symptoms onset. In addition, we have shown that these rapid immunoassays can be used to infer the neutralizing activity of sera against SARS-CoV-2.

## Data Availability

The authors confirm that the data supporting the findings of this study are available within the article [and/or] its supplementary materials.

## Acknowledgments

The authors would like to thank Gert Zimmer (Institute of Virology and Immunology, Mittelhäusern/Switzerland), Stefan Pöhlmann and Markus Hoffmann (both German Primate Center, Infection Biology Unit, Goettingen/Germany) for providing the reagents required for VSV pseudotype generation. Eliseo Albert holds a Río Hortega research contract from the Carlos III Health Institute (Ref. CM18/00221). Ron Geller holds a Ramón y Cajal fellowship from the Spanish Ministry of Economy and Competitiveness (RYC-2015-17517).

## Compliance with ethical standards

### Conflict of interest

The authors have no conflict of interest.

### Ethical statement

The current study was approved by the Research Ethics Committee of Hospital Clínico Universitario INCLIVA (March, 2020).

### Informed consent

Not applicable (as discussed with the institutional medical Ethical committee).

### Funding

This work was supported by Valencian Government grant IDIFEDER/2018/056 to JRD, Generalitat Valenciana grant Covid_19-SCI to RG-R, Spanish National Research Council grant CSIC-COV19-082 and Fondo Supera Covid-19 grant BlockAce to RG-R.

### Author’s contribution

AV: Methodology, data analysis, validation, review & editing; IT: Formal analysis, review & editing; VL: Methodology, investigation; CFG: Methodology, investigation; EA: Resources, project administration, review & editing; RG-R: Methodology, investigation, validation, funding acquisition, review & editing; MJA: Methodology, data analysis, validation, review & editing; JB: Supervision; review & editing; EG, Data analysis, validation, review & editing; JRD: Conceptualization, supervision, funding acquisition, review & editing; RG: Methodology, investigation, validation, funding acquisition, review & editing; DN: Conceptualization, supervision, writing the original draft, review & editing.

## Notes

### Competing Interest Statement

The authors have declared no competing interest.

### Author Declarations

The current study was approved by the Research Ethics Committee of Hospital Clinico Universitario INCLIVA (March, 2020).

